# Sub-diagnostic effects of genetic variants associated with autism

**DOI:** 10.1101/2021.02.12.21251621

**Authors:** Thomas Rolland, Freddy Cliquet, Richard J.L. Anney, Clara Moreau, Nicolas Traut, Alexandre Mathieu, Guillaume Huguet, Jinjie Duan, Varun Warrier, Swan Portalier, Louise Dry, Claire S. Leblond, Elise Douard, Frédérique Amsellem, Simon Malesys, Anna Maruani, Roberto Toro, Anders D. Børglum, Jakob Grove, Simon Baron-Cohen, Alan Packer, Wendy K. Chung, Sébastien Jacquemont, Richard Delorme, Thomas Bourgeron

## Abstract

While over a hundred genes are significantly associated with autism, little is known about the prevalence of variants affecting them in the general population. Nor do we fully appreciate the phenotypic diversity beyond the formal autism diagnosis. Using data from more than 13,000 autistic individuals and 210,000 undiagnosed individuals, we provide a gene-level map of the odds ratio for autism associated to rare loss-of-function (LoF) variants in 185 genes robustly associated with autism, alongside 2,492 genes displaying intolerance to LoF variants. In contrast to autism-centric approaches, we investigated the phenotype of undiagnosed individuals heterozygous for such variants and show that they exhibit a decrease in fluid intelligence, qualification level and income, and an increase in material deprivation. These effects were larger for LoFs in autism-associated genes than in other LoF-intolerant genes and appeared largely independent of sex and polygenic scores for autism. Using brain imaging data from 21,049 UK-Biobank individuals, we provide evidence for smaller cortical surface area and volume among carriers of LoFs in genes with high odds ratios for autism. Our gene-level map is a key resource to distinguish genes with high and low odds ratio for autism, and highlights the importance of including quantitative data on both diagnosed and undiagnosed individuals to better delineate the effect of genetic variants beyond the categorical diagnosis. Data are available at https://genetrek.pasteur.fr/.

## MAIN

Autism is a heterogeneous condition characterized by atypical social communication, as well as unusually restricted or stereotyped interests^1^. Its genetic architecture ranges from monogenic (for example caused by a *de novo* variant) to complex polygenic forms, i.e. driven by the cumulative effect of a multiple common variants, each having a small effect^2^. In the past 20 years, there has been tremendous progress in identifying genes robustly associated with autism^3,4^, and more widely with neurodevelopmental disorders (NDD)^5–7^, including intellectual disability (ID), delayed developmental milestones, and epilepsy^8,9^.

Little is known about the prevalence of rare loss-of-function (LoF) variants within these genes in the general population. Nor do we understand the inter-individual phenotypic variability of carriers beyond the autism diagnosis^10,11^. In this study, we analyzed whole-exome sequencing (WES) data from four studies, for a total of 226,664 individuals of European descent (Extended Data Fig. 1 and Methods); 13,091 individuals diagnosed with autism (henceforth, autistic individuals), 19,488 first-degree relatives of autistic individuals and 194,085 individuals identified from unselected population samples (Extended Data Fig. 2 and Methods).

First, we listed a set of 185 autosomal genes with dominant mode of inheritance, that are more frequently mutated in autistic individuals than in control individuals (Extended Data Table 1, Methods). We note here that despite no evidence linking these genes specifically to autism compared to other neurodevelopmental conditions^5–7^, we refer to these genes in this study as “autism-associated genes”. In addition, we analyzed 2,492 genes not considered as autism-associated genes, but with evidence for intolerance to LoF variants in reference populations (hereafter referred to as “constrained genes”, Extended Data Table 2, Methods)^12^.

Second, we identified high-confidence rare LoF variants (frequency less than 1% in each study) that were absent from the reference European population in the Genome Annotation Database (gnomAD; https://gnomad.broadinstitute.org/)^12^. We focused this study on LoF variants since 85% of known autism-associated genes are considered as intolerant to LoF variants (Extended Data Fig. 3)^12^. Because the impact of a LoF variant might depend on its location in the coding region^12,13^, we further selected stringent LoF variants (S-LoFs) that fell in an exon retained in more than 10% of the brain transcripts of the corresponding gene and truncating more than 10% the encoded protein (Methods). We observed S-LoFs in autism-associated genes in 4% of autistic individuals (n=524, 95% confidence interval [3.67%-4.34%]), 1.15% of their siblings and parents (n=224, 95% CI [1%-1.3%]), and 0.61% of individuals from UK-Biobank (n=1,161, 95% CI [0.58%-0.65%], Fig. 1a). We also observed that 35% of the S-LoFs in autism-associated genes identified among undiagnosed individuals fall within the same exons as those identified among autistic individuals (Extended Data Fig. 4), suggesting that these variants should have very similar consequences on the encoded protein independently of the status of the individuals^14^.

**Figure 1.**
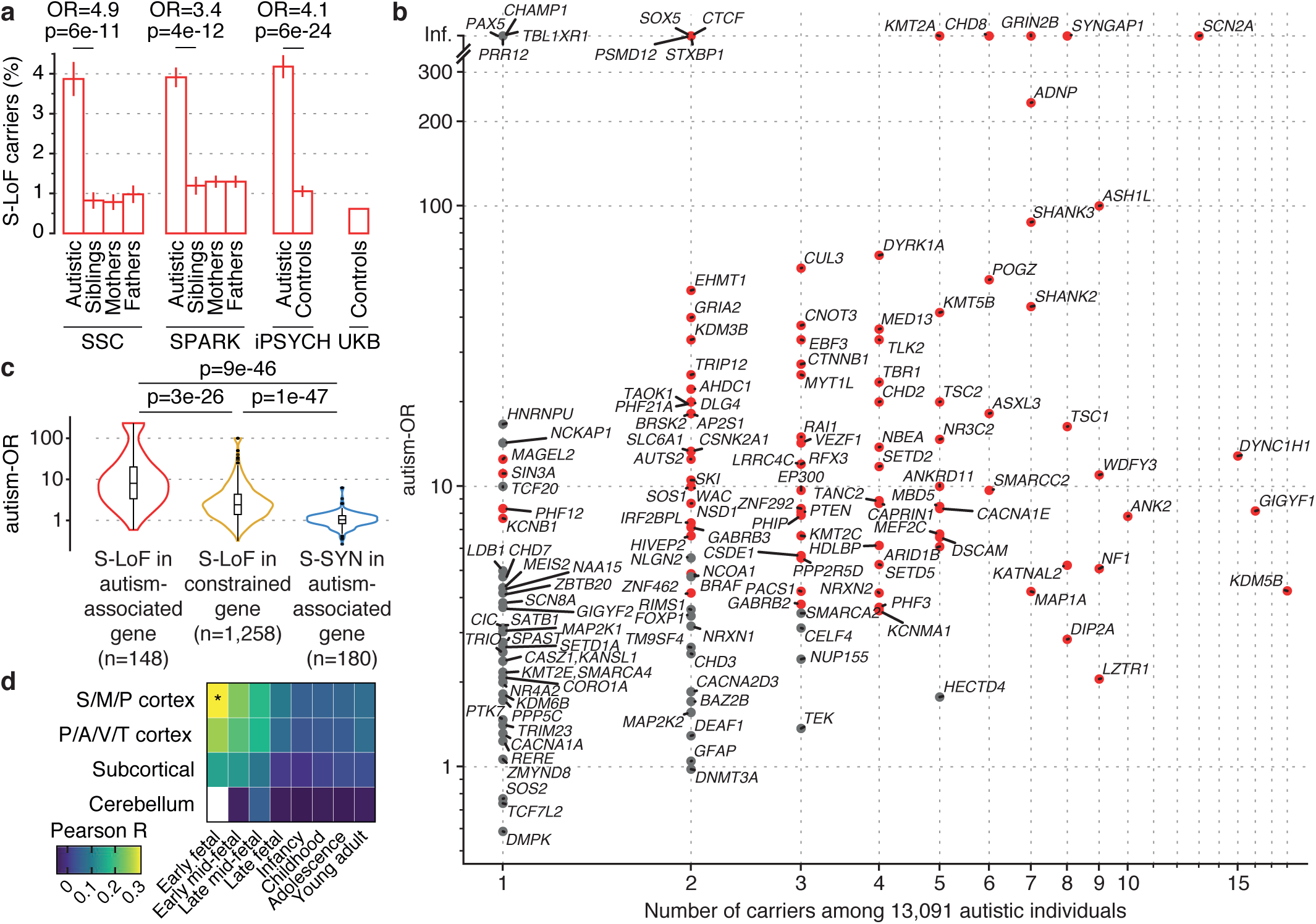
Gene-level autism odds ratio for rare variants in autism-associated and constrained genes. (**a**) Fraction of individuals carrying high-confidence rare stringent loss-of-function variants (S-LoFs) in autism-associated genes in each sample, stratified by status and family relationship. Error bars correspond to standard errors of the proportions. Odds ratios and p-values from two-sided Fisher exact tests comparing autistic children and their siblings in SSC and SPARK samples, and autistic and control individuals in the iPSYCH sample. SSC: Simons Simplex Collection, SPARK: Simons Powering Autism Research for Knowledge, iPSYCH: The Lundbeck Foundation Initiative for Integrative Psychiatric Research, UKB: UK-Biobank. (**b**) Number of S-LoF carriers among autistic individuals and autism-OR, e.g. enrichment in autistic individuals compared to undiagnosed individuals (based on 100 sub-samplings of undiagnosed individuals to match the number of autistic individuals, see Methods). Genes with autism-ORs significantly higher than expected by chance (empirical test based on 10,000 bootstraps, see Methods) are shown in red, others in grey. (**c**) Distribution of gene-level autism-OR of S-LoFs in autism-associated genes, S-LoFs in constrained genes and S-SYNs in autism-associated genes. P-values measured using two-sided Mann-Whitney U-tests, corrected for multiple testing using the Bonferroni method. (**d**) Correlation between autism-OR and gene expression in distinct brain regions and developmental periods for 130 genes for which at least one variant was identified among autistic individuals (expression data for early fetal cerebellum was not available, see Methods). Cortical regions were grouped as follows: posterior inferior parietal cortex, primary auditory cortex, primary visual cortex, superior temporal cortex, inferior temporal cortex (P/A/V/T cortex); primary somatosensory cortex, primary motor cortex, orbital prefrontal cortex, dorsolateral prefrontal cortex, medial prefrontal cortex, ventrolateral prefrontal cortex (S/M/P cortex). Correlations and p-values measured by Pearson correlation tests between autism-OR and gene expression, and p-values were corrected for multiple-testing using the FDR method (*corrected p=0.048). For panels (**c**) and (**d**), we set infinite autism-OR values to the highest measurable autism-OR in the corresponding gene set.

We then estimated for each gene the odds ratio for autism (autism-OR) of S-LoFs (Fig. 1b), i.e. the difference in the number of carriers of S-LoFs among autistic and undiagnosed individuals, adjusting for the large difference in sample size between cases and controls. To improve the robustness of our estimations, we used a sub-sampling procedure and further selected singleton S-LoFs (Extended Data Fig. 5, Methods). Prevalence, autism-OR and aggregated variant data can be visualized and downloaded on https://genetrek.pasteur.fr/^15^. Several autism-associated genes such as *SCN2A, ASH1L* and *ANK2* were among the most frequently mutated genes in autistic individuals (Fig. 1b), but displayed distinct autism-ORs (e.g., *SCN2A*=Inf.; *ASH1L*=100.1; *ANK2*=7.8). *SCN2A* was among 13 autism-associated genes (Extended Data Table 3) such as *CHD8, GRIN2B* and *SYNGAP1* for which all variants identified in autistic individuals were found *de novo*^16^, and for which no carrier of S-LoFs were identified among the 213,573 undiagnosed individuals. In contrast, for 135 autism-associated genes including *ASH1L, ANK2* and *SHANK3*, we could identify at least one carrier of a S-LoF among the undiagnosed individuals, suggesting lower effect sizes on autism diagnosis (Fig. 1b, Extended Data Table 3). We observed that four genes (*AP1S1, GIGYF1, PTEN* and *SHANK2*) displayed an autism-ORs above 8 while they were considered as relatively tolerant to LoF variants in literature (Extended Data Table 1), supporting caution in applying specific cutoffs for LoF intolerance metrics^17^. Altogether our results indicated that an exhaustive investigation of less penetrant variations is warranted to complete the map of genes associated to autism and more generally to NDD^18,19^.

To compare the effect of S-LoFs in autism-associated genes with other types of variants and sets of genes, we subsequently measured the autism-OR of synonymous variants (S-SYNs) in autism-associated genes and of S-LoFs in 2,492 constrained genes (Extended Data Fig. 6 and Extended Data Tables 4 and 5). As expected, S-LoFs in autism-associated genes displayed higher autism-ORs compared to S-LoFs in constrained genes (corrected p=3e-26, Mann-Whitney U-test) and S-SYNs in autism-associated genes (corrected p=9e-46) (Fig. 1c). Notably, some constrained genes such as *AP2M1, ZMYND11* and *CACNG2* reported in individuals with ID, displayed autism-ORs above 10 without being formally associated with autism (Extended Data Table 4).

To investigate the relationship between biological functions and the autism-OR, we studied the expression level of autism-associated genes in four different human brain regions and at eight different developmental periods. We found that the autism-OR was positively correlated with gene expression in early fetal period of cortex development (Pearson r=0.27 and FDR-corrected p=0.048 in auditory, visual, parietal and temporal cortex at the early fetal period, Fig. 1d; Extended Data Tables 6 and 7 and Methods)^20^. We also observed that genes encoding proteins associated with synapse function/architecture tended to display higher autism-ORs compared to genes not encoding synaptic proteins (nominal p=0.03; Extended Data Fig. 7 and Extended Data Table 8).

Besides rare variants with large effect, common variants associated with autism have been identified through genome-wide associations studies (GWAS) and can be aggregated to estimate a polygenic score for autism (autism-PGS) for each individual (Extended Data Fig. 8, Methods)^2,21,22^. Using logistic regression models, we measured the independent and interaction effects on autism diagnosis associated to the S-LoFs and the autism-PGS for 27,415 autistic individuals. We distinguished S-LoFs in genes below and above a threshold of autism-OR of 10 to quantify their differential effect on the autism diagnosis. All independent associations of S-LoFs, autism-PGS and sex with autism status were significant (Fig. 2a; Extended Data Tables 9 and 10). The association of S-LoFs with autism status was higher for genes with high compared to low autism-OR, 1.9 to 2.6 times higher for S-LoFs in autism-associated genes than in constrained genes, and 3.7 to 13.7 times higher for S-LoFs in autism-associated genes than for autism-PGS (Fig. 2a). We replicated these results in an independent analysis of the iPSYCH sample (Extended Data Fig. 9, Extended Data Table 11 and Methods).

**Figure 2.**
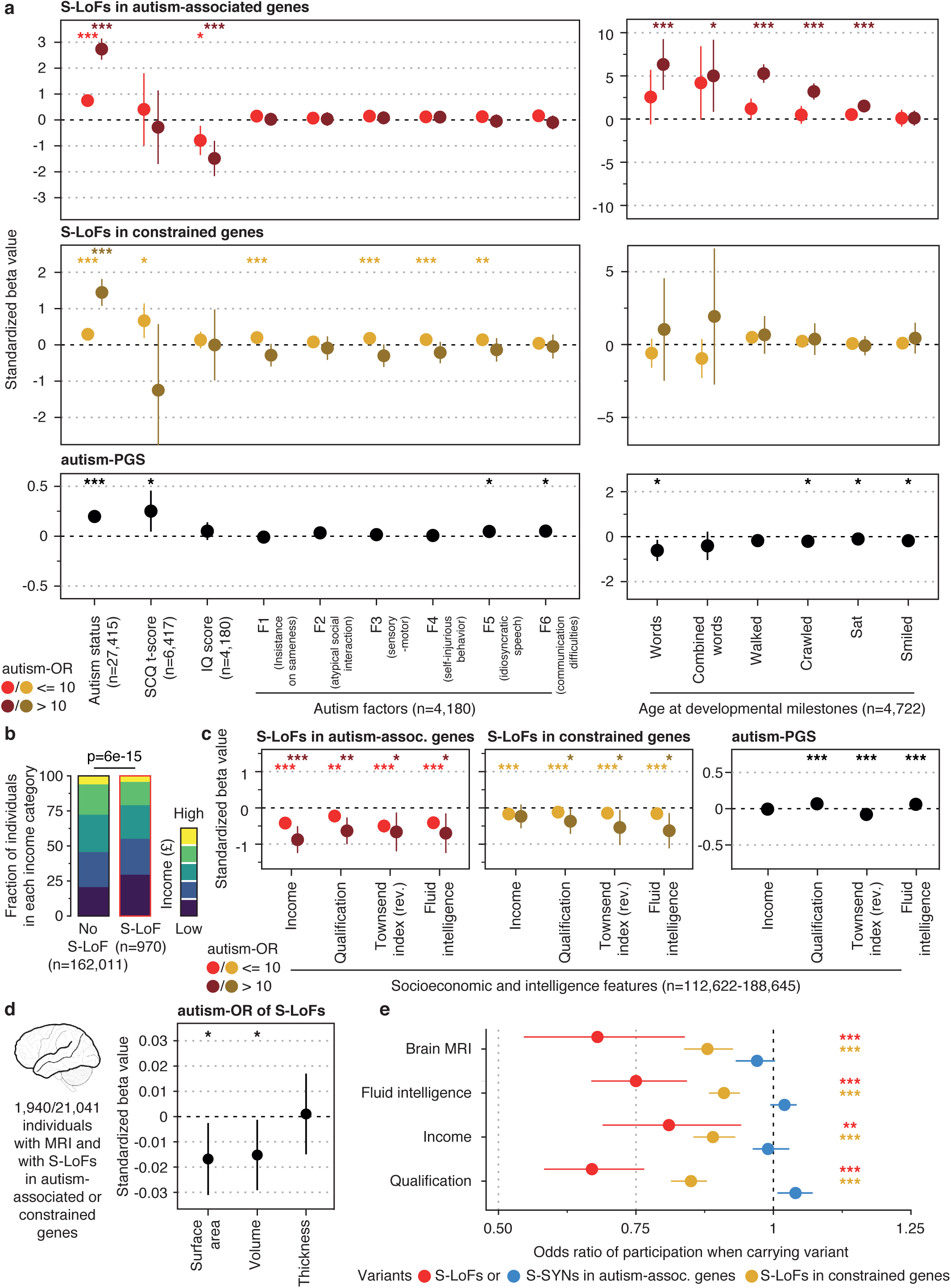
Sub-diagnostic effects of rare variants in autism-associated and constrained genes among diagnosed and undiagnosed individuals. (**a**) Standardized beta values associated to variant presence and autism-PGS from multivariable regression analyses of autism diagnosis, SCQ t-score, IQ score, autism factors and developmental milestones, stratified by gene type and autism-OR of genes carrying the variants (Methods). Regressions performed on individuals from the SSC and SPARK samples. To correct for the biased sex ratio among autistic individuals with approximately 1 girl for 4 boys, sex was added as an independent covariate. The beta values associated to autism-PGS when S-LoFs in constrained genes with autism-OR > 10 are considered in the regression analysis are shown (see Extended Data Tables 9 and 10 for complete results). Error bars correspond to 95% confidence interval. P-values associated with each beta value were corrected for multiple testing using the FDR method (***p<0.001, **p<0.01, *p<0.05). The number of individuals with available data is shown for each regression. For age at developmental milestones, higher values indicate higher age. (**b**) Distribution of incomes for carriers and non-carriers of S-LoFs in autism-associated genes among undiagnosed UK-Biobank individuals. The fraction of individuals in each category and the regression p-value for the association of variant presence with income change are reported. (**c**) Standardized beta values associated to variant presence and autism-PGS from multivariable regression analyses of socio-economic traits and fluid intelligence, stratified by gene type and autism-OR of genes carrying the variants (Methods). The Townsend index measures were reversed so that higher material deprivation was indicated with a negative sign. The number of individuals with available socio-economic or fluid intelligence data is shown. Legend as in (**a**). (**d**) Standardized beta values associated to autism-OR from multivariable regression analyses of brain anatomy among UK-Biobank undiagnosed individuals. Total values for cortical thickness, surface area and volume were measured as the sum of all 68 regions (Methods). S-LoFs in autism-associated and constrained genes were grouped to increase sample size (see Extended Data Table 12 for complete results). P-values were corrected for multiple testing using the FDR method (* corrected p-value = 0.048). (**e**) Odds ratio of participation when carrying a variant among UK-Biobank undiagnosed individuals for S-LoFs in autism-associated and constrained genes and for S-SYNs in autism-associated genes. P-values were corrected for multiple testing using the FDR method for each gene set and variant type independently (***p<0.001, **p<0.01).

We performed additional multivariable regression analyses to investigate the impact of S-LoFs and autism-PGS on several sub-diagnostic autism traits including age at developmental milestones, the social and communication questionnaire (SCQ) t-score, the IQ score, and six main autism-related factors previously described^6^ (F1: Insistence on sameness; F2: Atypical social interaction at age five; F3: Atypical sensory-motor behavior; F4: Self-injurious behavior; F5: Idiosyncratic repetitive speech and behavior; F6: Difficulties in communication) (Fig. 2a, Extended Data Tables 9 and 10). We observed a significant negative association of S-LoFs in autism-associated genes with IQ score and positive association with age at developmental milestones, but no association with SCQ t-score or autism-related factors (Fig. 2a), as previously reported for *de novo* variants among autistic children^23^. These associations showed a gradient of severity depending on the autism-OR (Fig. 2a). Notably, S-LoFs in constrained genes were mainly associated with non-social autism factors (F1, F3, F4 and F5) but not with SCQ t-score, IQ score and developmental milestones. The autism-PGS was associated with SCQ t-scores and factors related to difficulties in speech and communication (F5 and F6), suggesting an effect of the common variants on communication skills and repetitive speech/behaviors in autistic individuals. We finally observed no interaction between S-LoF and autism-PGS, suggesting mostly independent effects of rare and common variants on autism-related traits^21^.

We subsequently explored if, among undiagnosed UK-Biobank individuals, carriers of S-LoFs displayed differences in cognitive and socio-economical features compared to non-carriers. We performed multivariable regression analyses on fluid intelligence scores, qualification levels, income, and material deprivation estimated by the Townsend deprivation index (e.g., unemployment and non-home ownership) (Extended Data Tables 9 and 10). Individuals carrying S-LoFs in autism-associated genes displayed lower fluid intelligence, qualification, income (Fig. 2b) and higher material deprivation compared to non-carriers (Fig. 2c). These associations were stronger for S-LoFs in autism-associated genes than in constrained genes and showed a gradient of severity depending on the autism-OR. Interestingly, in contrast to the impact of S-LoFs, autism-PGS was positively associated with fluid intelligence and qualification level. However, as for S-LoFs, autism-PGS was also associated with increased level of the Townsend deprivation index (Fig. 2c). Altogether our results indicated that S-LoFs mostly affect the cognitive skills of individuals rather than their socio-communication abilities, as previously reported for large copy-number variants or *de novo* single nucleotide variants^6,24–26^.

Several autism-associated variants have been shown to modify brain structure^27–29^, and we finally questioned if S-LoFs had an impact on brain anatomy using magnetic resonance imaging (MRI) data from 21,041 UK-Biobank individuals. To increase our prediction power, we grouped the 1,940 carriers of S-LoFs in autism-associated or in constrained genes, and tested whether autism-OR was associated with differences in global cortical volume, thickness and surface area using multivariable regression analyses (Extended Data Table 12, Methods). We observed that the autism-OR was significantly associated with slight reduction of cortical surface area and volume (beta=-0.017 and beta=-0.015, respectively, corrected p=0.048 for both; Fig. 2d). We found no significant association of autism-OR with any specific brain region, suggesting that larger sample sizes will be required to further investigate the effect of S-LoFs on brain anatomy^30^.

UK-Biobank individuals are not a perfectly accurate representation of the general population^31^, and participation bias has a genetic component^32,33^. We observed a significant negative effect of S-LoFs on response to questionnaires exploring qualification level, income, and fluid intelligence (Fig. 2e, Extended Data Table 13, Methods). This effect was higher for S-LoFs in autism-associated genes than for constrained genes and was absent for S-SYNs in autism-associated genes. Participation to brain MRI scanning showed the same trend, suggesting that the imaging subsample also present a participation bias^34^. These results provide additional support that UK-Biobank sample may suffer from a “healthy volunteer bias”, which alter our ability to quantify the actual effect of genetic variants.

In summary, by systematically analyzing WES data of more than 13,000 autistic individuals and 210,000 undiagnosed individuals, we estimated the autism-OR of rare LoF variants in 185 genes associated with autism. As expected, the genes with the highest autism-ORs (e.g. *DYRK1A, GRIN2B, SCN2A* and *SYNGAP1)* were those repeatedly identified as affected by *de novo* variants in independent genetic studies of autism. The reasons why some individuals carrying the S-LoF will have a diagnosis of autism, and some do not, probably depends on additional genetic, epigenetic, and environmental factors. For example, the location of the variant can be critical^35^. We found three undiagnosed individuals who carried S-LoFs impacting *SHANK3* (Extended Data Fig. 4), but two of them affected the alpha isoform, which was known to be associated with milder phenotypes^36^ compared to other *SHANK3* isoforms^37^. Hence, in addition to a gene map, an exon or even site-specific map might be more accurate to assess the penetrance of the LoF variants^38^, but this level of accuracy will require even larger sample size cohorts.

Sex could also be a factor modulating the penetrance of genetic variants. For some specific genes or pathways, penetrance of genetic variants could be different in males and females^1,11,39^. For example, inherited variants in autosomal genes such as *SHANK1* have been reported to be more frequently transmitted by mothers and lead to autism preferentially or exclusively in males^40^. In our study, we observed a significant enrichment of female autistic individuals carrying S-LoFs in autism-associated genes compared to autistic males (odds ratio 1.71, p=5e-05, Fisher exact test) as previously reported^41,42^. This may imply that males can have an autism diagnosis through other types of genetic architectures (e.g. lower load of common variants compared to females)^6,43^. While our sample size was relatively large, it was not large enough to robustly investigate the gene-level autism-OR of S-LoFs for males and females independently (Extended Data Fig. 10). We did not observe overall differences in sex-ratio amongst non-autistic carriers of S-LoFs affecting autism-associated genes, as previously reported for parents of children with NDD^44^ or for non-autistic siblings^8,39^.

Finally, the genetic background could also modulate the penetrance of LoFs as recently reported in carriers of the 22q11 deletion in schizophrenia^45^. In our study, we observed significant independent effects of S-LoFs and autism-PGS on autism-related traits, but could not detect a significant interaction between them, suggesting these two genetic factors act independently and additively on autism liability^21^. However, we might be underpowered to detect such interaction^21^, especially if the interplay between rare and common variants diverges from one gene to another. Averaging the effect over multiple genes and individuals would tend to weaken the strength of the association. Integration of additional polygenic scores and data related to expression levels (expression quantitative trait loci, eQTL) is warranted to provide a better estimate of the phenotype of carriers^45–47^ and to enhance our understanding of the biological pathways associated with autism^22,48^. Epigenetic/environmental factors might also modulate the penetrance of the genetic variants, but large-scale data to detect their impact are lacking so far.

To conclude, we show that LoF variants in autism-associated genes do not result in a systematic clinical diagnosis of autism in individuals, but influence the global functioning of the carriers as indicated by cognitive and socio-economic metrics. Such fine-grained estimation of the autism-OR associated with variants in autism-associated genes has important consequence for clinical counseling as they support for the vast majority of the autism-associated genes a complex interplay between gene-level variations and clinical outcome^49,50^. Future large-scale studies integrating phenotypic information beyond diagnostic criteria should open the path towards the identification of genetic and environmental factors that could help individuals to cope with the adverse consequences of carrying such variations.

## Data Availability

Whole-exome and SNP genotyping data from the SSC and SPARK cohorts can be obtained by applying at SFARI Base (https://www.sfari.org/resource/sfari-base/). The UK-Biobank whole-exome, SNP genotyping and brain imaging data can be obtained by applying at the UK-Biobank database (https://www.ukbiobank.ac.uk/). The human neurodevelopmental transcriptome dataset is available on the BrainSpan database (http://www.brainspan.org). Functional annotations can be obtained from SynGO (https://syngoportal.org/) and Gene Ontology (http://current.geneontology.org/annotations/goa_human.gaf.gz). Electronic health records and healthcare claims data used in the present study for the UK-Biobank individuals are not publicly available due to patient privacy concerns. Prevalence and autism-OR measures can be visualized and downloaded on https://genetrek.pasteur.fr/.

http://www.brainspan.org

https://syngoportal.org/

http://current.geneontology.org/annotations/goa_human.gaf.gz

https://www.ukbiobank.ac.uk/

## METHODS

### Samples

For the SSC, SPARKv1 and SPARKv2 cohorts, we downloaded genetic and clinical data from SFARI Base (https://sfari.org/sfari-base). For the SSC cohort, we selected 10,141 individuals with both whole-exome sequencing (WES) and SNP array data, who were not twins and did not show high number of erroneous variant calls (families filtered out: 12958, 14572 and 11037). For the SPARKv1 cohort, we selected 19,671 individuals with both WES and SNP arrays data, who were not withdrawn, not twins and not showing excessive number of variants or abnormal age, and from families in which both parents were undiagnosed and had available genetic data. For the SPARKv2 cohort, we selected 5,970 individuals with both WES and SNP arrays data, who were not withdrawn, and from families in which both parents were undiagnosed and had available genetic data. For simplicity, the SPARKv1 and SPARKv2 samples were merged into a SPARK sample.

For the UK-Biobank cohort, we downloaded genetic, demographic and brain imaging data from the UK-Biobank database (projects 51869, 40980 and 18584). We selected 200,457 individuals with both WES and SNP arrays data, who were not twins (kinship < 0.4 from relationship file of UK-Biobank), and who did not report autism-related symptoms (based on ICD10-F84 index or the autism diagnostic questionnaire).

For the iPSYCH sample, we downloaded tabular files for each gene of interest from the ASC website (https://asc.broadinstitute.org), and calculated the maximum of the allele numbers per status for all variants, corresponding to 4,811 autistic individuals and 5,214 control individuals.

### Autism and constrained gene sets

We focused on coding exons of 220 autism-associated genes: genes from the SFARI Gene database with a score of 1 (https://gene.sfari.org/database/human-gene/),102 genes from a recent case-control study of rare variations^8^, and 157 genes robustly associated with autism in multiple independent studies and unrelated individuals by the SPARK committee (http://sparkforautism.org) (Extended Data Table 1).

Constrained genes were defined based on suggested thresholds of the LOEUF (loss-of-function observed/expected upper bound fraction < 0.35) or the pLI (probability of loss-of-function intolerance > 0.9), both extracted from the gnomAD website (https://gnomad.broadinstitute.org)^12^.

The current study focused on autosomal genes, and we filtered out the genes with an evidence of recessive type of inheritance^15^.

For sex-specific analyses of autism odds ratio, all autism-associated genes on the X chromosome were also considered for male-specific analyses, and only if they had no evidence of recessive type of inheritance for female-specific analyses (dominant: *ARHGEF9, CASK, CDKL5, DDX3X, FMR1, HNRNPH2, IQSEC2, MECP2, NEXMIF, PCDH19, USP9X*; recessive: *AFF2, ARX, ATRX, KDM5C, NLGN3, NLGN4X, PTCHD1, SLC9A6, SYN1, UPF3B*).

### SNP arrays

For the SSC sample, the GRCh36-based SNP array data for the 3 different technologies (Illumina Omni1Mv1, n = 1,354, Omni1Mv3, n = 4,626, and Omni2.5, n = 4,240) were downloaded from the SFARI Base (https://sfari.org/sfari-base), and 15 individuals were removed because they were twins. Arrays from each technology were mapped onto the GRCh37 human genome version separately. We downloaded the pre-processed GRCh37-based genotyping files of 26,879 SPARKv1 and 15,904 SPARKv2 participants from the SFARI Base. SSC and SPARK genotyping files were filtered from ambiguous SNPs (A/T & G/C SNPs if MAF > 0.4, SNPs with differing alleles, SNPs with > 0.2 allele frequency difference, SNPs not in reference panel) and imputed on the HRC panel version r1.1^51^ on the Michigan servers with default parameters^52^. GRCh37-based imputed genotyping files for 200,080 UK-Biobank individuals were downloaded from the UK-Biobank database (projects 51869 and 18584). After imputation we kept only variants with a r2>=0.8 and merged the 3 different SNP arrays technologies from the SSC sample keeping only SNPs shared between all 3 technologies.

### Admixture

We used the 1000genome sequencing data of 2,504 individuals as a reference group of individuals of known ancestry^53^. We selected the 1000g SNPs that were present in the SSC, SPARKv1 and SPARKv2 datasets to perform a combined admixture for SFARI-base samples, and 1000g SNPs that were present in the UK-Biobank dataset to perform a separate admixture, using the Admixture v1.3.0 tool^54^ on 1 to 8 clusters. SSC, SPARKv1 and SPARKv2 genotypes, as well as UK-Biobank genotypes, were projected on the corresponding admixture models based on 1000g data, and we selected 5 clusters for separating the individuals by ancestry, corresponding to a low cross-validation error in both admixture models (Extended Data Fig. 1). Based on the reference EUR super-population, we used a fraction of each individual’s SNPs predicted as European ancestry threshold of >= 60% to define individuals as being of European ancestry, resulting in 8067, 15,360, 4,346 and 188,880 individuals in SSC, SPARKv1, SPARKv2 and UK-Biobank samples, respectively.

### Whole exome sequences

We downloaded the GRCh37 aligned BAM files of 8,960 SSC participants from SFARI Base (https://sfari.org/sfari-base). We then called the variants using GATK 3.8 following the BROAD Institute Best Practices^55^, and lifted over all variants to the GRCh38 human genome version. We downloaded the preprocessed GRCh38-based pVCF files of 27,270 SPARKv1 and 16,004 SPARKv2 participants from SFARI Base. All functional-equivalent (FE) GRCh38-based pVCF files for 200,642 UK-Biobank participants were downloaded from the UK-Biobank database (projects 51869 and 18584). All variants from SSC, SPARK and UK-Biobank samples were filtered for call rate > 0.9, genotype quality >= 30, depth > 20, allelic fraction >= 0.25 (and <= 0.75 for autosomal variants). Tabular lists of variants from the iPSYCH individuals were downloaded from the ASC website (https://asc.broadinstitute.org), and mapped to the GRCh38 human genome version (using chain file hg19toHg38.over.chain.gz).

We used VEP^56^ (using Ensembl 101) to annotate the variants. Non-neuro (individuals that were not cases of a few particular neurological disorders) non-Finnish European population frequencies were extracted using gnomAD exomes r2.1.1^12^. Variants with a MAF > 1%, present in > 1% of each sample or affecting genes that were recurrently found mutated across different individuals in different families (*MUC4, MUC12, HLA-A, HLA-B, HYDIN, TTN, PAX5, OR2T10, MYH4*) were filtered out. We used Loftee^12^ to filter low-confidence variants or variants corresponding to ancestral alleles, as well as variants annotated with any flag by Loftee. All LoF variants affecting autism-associated genes were visually validated with IGV^57^ on BAM/CRAM files for SSC, SPARK and UK-Biobank samples.

For the independent regression analyses on autism status in the iPSYCH sample, we performed additional quality control steps on the 236 S-LoFs in autism-associated genes and 1,345 S-LoFs in constrained genes. The initial QC steps for the iPSYCH Danish Blood Spot whole-exome sequencing data has been described previously^58^. Briefly, after first round sample-level and variant-level QC, three call rate filters were used subsequently, 1) remove variants with a call rate < 90%, 2) remove samples with a call rate < 95%, and 3) remove variants with a call rate < 95%. Between the sample call rate filter and the final variant call rate filter, one of each pair of related samples (relatedness as a pi-hat value ≥ 0.2) was removed. Subsequently, we selected for this study the cases diagnosed with autism no later than by the end of 2016. This gave us a study sample of 4,622 cases and 4,753 controls. We defined rare variants as with an allele count no greater than five across our dataset (n=9,375) and the non-Finnish Europeans from non-psychiatric exome subset of the gnomAD (n=44,779). We matched these S-LoFs to the original S-LoFs and identified 138 out of 236 S-LoFs in autism-associated genes and 767 out of 1,345 S-LoFs in constrained genes in iPSYCH. Replication analyses were based on these S-LoFs.

### Relative position on encoded protein and pext score

We annotated the relative position of the variants on the encoded protein using the Loftee coding sequence (CDS) position when available or VEP CDS position otherwise, and the CDS size for each transcript from BioMart^59^. To measure exon usage in different isoforms of each gene within brain tissues, we downloaded the base-level pext score from the gnomAD website (https://gnomad.broadinstitute.org)^13^. Briefly, the pext score summarizes the isoform expression values across tissues and allows to measure the expression status of exonic regions across tissues, at the exon level. For each exon of each gene, we selected the maximum value of the pext measures from 13 brain tissues (amygdala, anterior cingulate cortex BA24, caudate basal ganglia, cerebellar hemisphere, cerebellum, cortex, frontal cortex BA9, hippocampus, hypothalamus, nucleus accumbens basal ganglia, putamen basal ganglia, spinal cord, substantia nigra). For splice-site variants, we measured the relative position and pext score based on the closest coding exon (position of the variant +/- 3 bp). We finally filtered variants using the pext score, reflecting how much the corresponding exon was expressed in brain tissues.

### Gene level autism odds ratio

The odds ratio for autism (autism-OR) was measured to estimate the strength of the association between outcome (autism diagnostic) and genetic risk factors (carrying a LoF variant) for each gene, using the following formula:

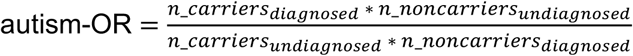

Given the large difference in sample size between diagnosed and undiagnosed individuals and given that the definition of rarity of variants depends on the sample size, we performed 100 iterations of a sub-sampling procedure: 1) randomly selecting as many undiagnosed individuals as diagnosed individuals, and 2) selecting singletons among diagnosed individuals and among undiagnosed individuals separately. We then used the average number of carriers among undiagnosed individuals to estimate the autism-OR for each gene. To compare the autism-OR to what would be expected by chance given our samples, we also performed a bootstrapping procedure, randomly selecting as many individuals as diagnosed individuals, artificially labelling them as diagnosed and labelling the rest of the sample as undiagnosed, and measuring the autism-OR using the same algorithm. We ran this procedure 10,000 times, measured for each gene the number of times M the expected autism-OR was higher or equal to the observed autism-OR, divided it by the number of bootstraps performed N and used the (M+1)/(N+1) ratio as an empirical p-value. The 95% confidence interval around this empirical p-value was measured using the following formula to assess the degree of certainty of the empirical p-value.

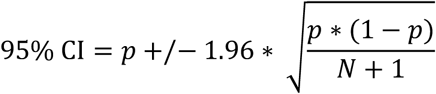

We verified that all reported signals for the analyses described in Fig. 2 were similar when restricting the analyses to genes with autism-ORs significantly higher than expected by chance (upper fraction of the 95% CI of the empirical p-value < 0.05), with the exception of the significance of the brain anatomy results that were insufficiently powered.

### Developmental brain gene expression and enrichment in autism pathways

The developmental brain transcriptome data from 42 specimen and up to 16 brain structures was downloaded from the Allen Brain Atlas BrainSpan database (https://www.brainspan.org/). Only expression RPKM values (Reads Per Kilobase of exon model per Million mapped reads) above 1 were considered for expression analysis. Values for each gene were averaged across four brain regions and eight developmental periods as previously described^20^. Brain regions were defined as follows: R1) posterior inferior parietal cortex, primary auditory cortex, primary visual cortex, superior temporal cortex, inferior temporal cortex; R2) primary somatosensory cortex, primary motor cortex, orbital prefrontal cortex, dorsolateral prefrontal cortex, medial prefrontal cortex, ventrolateral prefrontal cortex; R3) striatum, hippocampus, amygdala; R4) mediodorsal nucleus of the thalamus, cerebella cortex. Developmental periods were defined as follows: P1) early fetal; P2) early mid-fetal; P3) late mid-fetal; P4) late fetal; P5) infancy; P6) childhood; P7) adolescence; P8) Young adult. Note that only one individual was available for P1R4 in the BrainSpan database, the corresponding period/region was therefore not investigated in this study. For the analysis of the correlation between gene expression and autism-OR, we artificially replaced infinite autism-OR values by the highest measurable autism-OR in the gene set, and the Pearson correlation test was performed in the log10 space for both expression and odds ratio of autism-associated genes. We verified that Kendall and Spearman rank-based correlations showed the same trend (data not shown).

Functional annotation of synaptic proteins were taken from SynGO^60^ and transcription proteins extracted from Gene Ontology term Transcription, DNA templated^61^.

### Autism polygenic score computation

SSC, SPARKv1, SPARKv2 and UK-Biobank imputed genotyping data were filtered separately from variants absent from more than 1% of individuals (geno001 parameter), then variants present in all 4 samples were merged with PLINK 1.9^62^. The polygenic score (PGS) for autism was computed by using the GWAS summary statistics from the Integrative Psychiatric Research Consortium (iPSYCH) and the Psychiatric Genomics Consortium (PGC)^2^. To exclude overlap in participants from the test and discovery data in the PGS analysis, the GWAS meta-analysis summary statistics reported^2^ were recalculated with the SSC data excluded. We used the SBayesR^63^ method of the GCTB tool v2.02 with the banded linkage disequilibrium (LD) matrix^64^ and suggested options (https://cnsgenomics.com/software/gctb) on the PGC-ASD summary statistics to estimate the posterior statistics of SNP effects. We finally computed the autism-PGS using PLINK 1.9 based on SBayesR-derived statistics for common SNPs (MAF > 10%).

We performed a PCA analysis using PLINK 2.0, and extracted the four first principal components to control for population structure when using the autism-PGS in regression analyses.

### Psychiatric, developmental, cognitive and socioeconomic features

The Social Communication Questionnaire results for SSC and SPARK samples were downloaded from SFARI Base (https://sfari.org/sfari-base) and were available for 8,235 probands and 4,176 non-autistic siblings of European ancestry. The autism factors and IQ score for SSC and SPARK samples were available for 4,180 probands from a previous study^6^. The developmental milestones for SPARK samples were downloaded from SFARI Base (https://sfari.org/sfari-base) and were available for 4,722 probands.

For the UK-Biobank individuals, age when attending assessment center and genetic sex were available for all 188,880 control European individuals. The Fluid Intelligence Test of reasoning and problem solving were completed by 112,626 individuals. We used the highest qualification an individual had achieved (e.g. university/college degree, A-levels), excluded participants with only ‘other professional qualifications’ and those who did not provide an answer to this question, retaining data for 146,864 individuals and categorizing in five bands (CSEs or equivalent, O levels/GCSEs or equivalent, NVQ or HND or HNC or equivalent, A levels/AS levels or equivalent and College or University degree). Annual income was categorized by the UK Biobank sample in five bands (<£18,000, £18,000–30,999, £31,000– 51,999, £52,000–100,000 and >£100,000), and was available for 162,987 participants. The Townsend Deprivation index of social deprivation was available on 188,654 individuals.

For participation analyses of qualification level, we considered as respondent participants who answered ‘other professional qualifications’, ‘CSEs or equivalent’, ‘O levels/GCSEs or equivalent’, ‘NVQ or HND or HNC or equivalent’, ‘A levels/AS levels or equivalent’ or ‘College or University degree’. For participation analyses of income, we considered as respondent participants who answered ‘<£18,000’, ‘£18,000–30,999’, ‘£31,000–51,999’, ‘£52,000–100,000’ and ‘>£100,000’.

### Brain structural anatomy

Imaging derived phenotypes (IDP) data were downloaded from the UK-Biobank database (projects 40980 and 18584). A total of 68 metrics for cortical regions, calculated using FreeSurfer and FSL software using the Desikan-Killiany Atlas^65^, were provided for 21,041 individuals with genetic data. Details of the acquisition protocol and imaging processing toolbox are available on the UK Biobank website: https://biobank.ctsu.ox.ac.uk/crystal/crystal/docs/brain_mri.pdf. Three global IDPs were investigated: total cortical volume, total cortical thickness and total cortical surface area. The total brain IDPs were obtained by summing left and right hemisphere global measures.

### Multivariable regression analyses

We performed ordinal logistic regression analyses for autism status using the following formula.

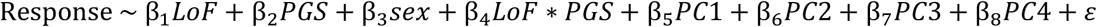

We performed linear regression analyses for SCQ t-score, IQ score, autism factors and developmental milestones on autistic individuals using the following formula.

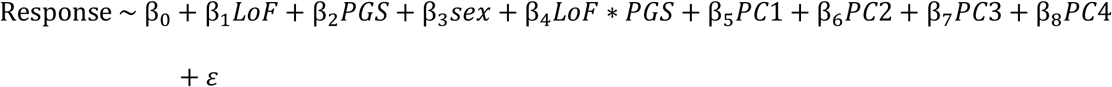

We performed linear regression analyses for fluid intelligence score and Townsend Deprivation index, and ordinal logistic regression analyses for income and qualification level on UK-Biobank individuals. For ease of interpretation, the directions of the Townsend Deprivation index effects were adjusted so that a negative sign always implied worse outcomes, e.g. higher Townsend Deprivation index.

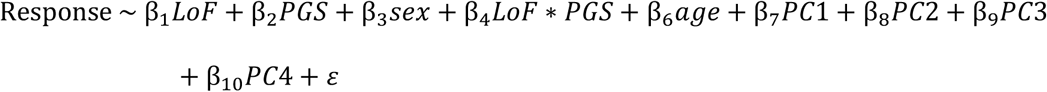

For brain anatomy among UK-Biobank individuals, multivariable linear regressions were performed separately for global cortical thickness, surface area and volume z-scored IDPs with the following formula, with the site variable representing the location where the scan was performed.

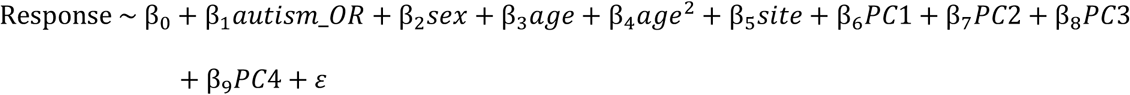

Multivariable linear regressions were performed separately for each 68 cortical regions using the following formula, adding the total measure for each metric (e.g. global cortical volume for the volume of the 68 cortical regions) as a covariate.

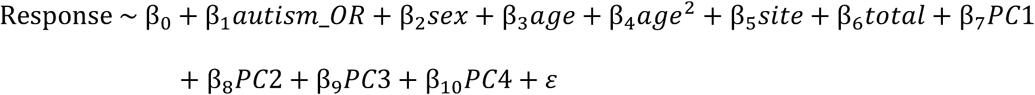

For all models, PC1 to 4 represent the first four principal components of the PCA based on genotyping data. Results were presented as standardized beta coefficients. To evaluate the significance of results, we used the Benjamini–Hochberg false discovery rate (FDR) method for p-value correction. Multiple-testing correction was applied separately for each covariate and independently for i) autism status, SCQ t-score, IQ score and autism factors, ii) developmental milestones, and iii) socioeconomic and fluid intelligence features. For multivariable analyses of brain anatomy, multiple-testing correction was applied separately for i) global measures (global cortical volume, surface area and thickness), and ii) cortical volume, iii) cortical thickness and iv) cortical surface area for region-level analyses.

### Statistical analyses

Most of the statistical analyses in this work were performed using statistical test implementations from python libraries scipy^66^ and statsmodels^67^. If not otherwise stated, analyses including adjusting p-values for multiple testing used the Benjamini and Hochberg control for false discovery rate^68^.

## CODE AVAILABILITY

Code used to implement the post-processing analyses in this paper is available at https://github.com/thomas-rolland/subdiagnostic-autism-variants.

## ACKNOWLEDGEMENTS

This research has been conducted using the Simons Simplex Collection and Simons Powering Autism Research for Knowledge from the Simons Foundation Autism Research Initiative, and the UK-Biobank cohort (projects 51869, 40980 and 18584). This work was supported by a grant from SFARI (#: 240059, TB). We are grateful to all of the families at the participating Simons Simplex Collection (SSC) sites, at the participating Simons Searchlight sites, the Simons Searchlight Consortium, as well as the principal investigators (A. Beaudet, R. Bernier, J. Constantino, E. Cook, E. Fombonne, D. Geschwind, R. Goin-Kochel, E. Hanson, D. Grice, A. Klin, D. Ledbetter, C. Lord, C. Martin, D. Martin, R. Maxim, J. Miles, O. Ousley, K. Pelphrey, B. Peterson, J. Piggot, C. Saulnier, M. State, W. Stone, J. Sutcliffe, C. Walsh, Z. Warren, E. Wijsman). We appreciate obtaining access to SNP arrays, WES and phenotypic data on SFARI Base. Approved researchers can obtain the SSC population dataset and the Simons Searchlight population dataset described in this study by applying at https://base.sfari.org. The authors would like to thank the members of the Human Genetics and Cognitive Functions lab for helpful discussions, and Kuldeep Kumar, Annabelle Harvey, Andréanne Proulx and Hanad Sharmarke for helping with the quality control of the UK-Biobank rs-fMRI preprocessed data. This work was funded by Institut Pasteur, the Bettencourt-Schueller Foundation, Université de Paris, the Conny-Maeva Charitable Foundation, the Cognacq Jay Foundation, the Eranet-Neuron (ALTRUISM), the GenMed Labex, AIMS-2-TRIALS which received support from the Innovative Medicines Initiative 2 Joint Undertaking under grant agreement No 777394 and the Inception program (Investissement d’Avenir grant ANR-16-CONV-0005). This project has received funding from the European Union’s Horizon 2020 research and innovative program CANDY under grant agreement No 847818. The views expressed here are the responsibility of the author(s) only. The EU Commission takes no responsibility for any use made of the information set out. The iPSYCH team was supported by grants from the Lundbeck Foundation (R102-A9118, R155-2014-1724, and R248-2017-2003) and the Universities and University Hospitals of Aarhus and Copenhagen. High-performance computer capacity for handling and statistical analysis of iPSYCH data on the GenomeDK HPC facility was provided by the Center for Genomics and Personalized Medicine and the Centre for Integrative Sequencing, iSEQ, Aarhus University, Denmark (grant to ADB). SBC received funding from the Wellcome Trust 214322\Z\18\Z, support from the European Union’s Horizon 2020 research and innovation programme and EFPIA and AUTISM SPEAKS, Autistica, SFARI. SBC also received funding from the Autism Centre of Excellence, SFARI, the Templeton World Charitable Fund, the MRC, and the NIHR Cambridge Biomedical Research Centre. The research was supported by the National Institute for Health Research (NIHR) Applied Research Collaboration East of England. Any views expressed are those of the author(s) and not necessarily those of the funder.

## AUTHOR CONTRIBUTIONS

T.R. and T.B. designed the research. T.R. performed all analyses, with the help of F.C., R.J.L.A., Al.M., G.H., J.D., V.W., C.S.L., E.D., A.D.B., J.G., S.B-C., A.P., W.K.C., S.J. and T.B. for the genomic analyses, C.M., N.T., S.P., L.D. and R.T for the analysis of brain imaging, and F.A., An.M., and R.D. for the clinical analyses. S.M. developed the website. T.R. and T.B. wrote the manuscript, with assistance from all other authors. All authors approved the manuscript.

## EXTENDED DATA FIGURES LEGENDS

**Extended Data Figure 1. Admixture results for UK-Biobank, SSC and SPARK individuals**. For each cohort, the cross-validation errors were shown for increasing values of clusters, and the resulting fraction of ancestry predicted in each admixture group was shown for five clusters. SSC and SPARK individuals were merged for this analysis. Fractions were shown for reference population individuals and for individuals of each cohort. The predicted European group, used for subsequent prediction of European ancestry (individuals with > 60% predicted fraction of European ancestry were considered European, see Methods), was shown in dark blue.

**Extended Data Figure 2. Framework of the study**. The schematic represents the different analyses performed in the manuscript, on all or subsets of the 13,091 diagnosed and 213,573 undiagnosed individuals with both whole-exome sequencing data available. Brain imaging and cognitive and functioning data were available only for a subset of the UK-Biobank individuals. SCQ t-scores, IQ scores, autism factors and developmental milestones were extracted only for a subset of diagnosed individuals from the SSC and SPARK samples. SNP array data were available for all individuals except from the iPSYCH cohort, which was treated separately (Methods).

**Extended Data Figure 3. Comparison of LoF and missense deleteriousness scores for autism-associated genes**. The suggested LOEUF threshold of 0.35, pLI threshold of 0.9 and top decile of the missense z-score distribution, corresponding to a value of 2.45, are shown.

**Extended Data Figure 4. Examples of variants mapping to exons of autism-associated genes**. For *SHANK3* and *NRXN2*, the variants identified in diagnosed and undiagnosed individuals are indicated on the Genetrek website. The four tracks correspond to variants identified in 13,091 autistic individuals, 194,085 control individuals, 15,200 parents of autistic individuals, and 4,288 non-autistic siblings of autistic individuals. For each track, the first line represents the variants identified, colored by variant type (S-LoF in autism-associated gene in red, S-LoF in constrained gene in orange, S-SYN in autism-associated gene in blue). The four other lines inform about the sex and inheritance mode of the identified variant (order: de novo in female, not de novo - “other” - in female, de novo in male, other in male), with grey marks corresponding to homozygous reference allele and blue marks heterozygous allele.

**Extended Data Figure 5. Gene-level autism-OR as a function of number of carrying individuals or families, average pext in brain tissues and relative position in encoded protein**. Fraction of individuals carrying S-LoFs, stratified by autism status, and corresponding gene-level autism-ORs as a function of thresholds in pext score, relative position on encoded protein and number of individuals or families. The fraction of undiagnosed individuals carrying S-LoFs corresponds to the average fraction of individuals in the 100 sub-sampling (Methods). The thresholds correspond to S-LoFs that were present in more than 10% of the brain-expressed transcripts, truncating more than 10% of the encoded protein, i.e. not in the last 10% of the protein sequence, and/or found in only one family or individual. The number of genes for which we find at least one diagnosed individual carrying a variant is indicated. P-values from Mann-Whitney U-tests.

**Extended Data Figure 6. Proportion of individuals carrying S-LoFs in autism-associated genes, S-LoFs in constrained genes or S-SYNs in autism-associated genes**. Fractions are shown in each sample, stratified by status and family relationship. Odds ratios (OR) and p-values from two-sided Fisher exact tests. Error bars correspond to standard errors of the proportions.

**Extended Data Figure 7. Biological pathways associated to high autism-OR**. Distribution of autism-OR for genes encoding synaptic and transcription proteins compared to autism-OR of genes not encoding such proteins. P-values from two-sided Mann-Whitney U tests.

**Extended Data Figure 8. Calculation of the autism-PGS**. (**a**) Nagelkerke’s pseudo-R2 correlation coefficient for the autism-PGS comparing autistic individuals and UK-Biobank individuals. R2 and p-value from a likelihood ratio test comparing the model including the autism-PGS to the null model including only the first four principal components of the PCA based on genotyping data. (**b**) Density plots of autism-PGS scores for autistic and UK-Biobank individuals.

**Extended Data Figure 9. Regression analysis for the effect of S-LoFs and autism-PGS on autism status in the iPSYCH sample**. Standardized beta values associated to variant presence and autism-PGS from multivariable regression analyses of autism status (Methods). The beta values associated to autism-PGS when S-LoFs in constrained genes with autism-OR > 10 are considered in the regression analysis are shown. Error bars correspond to 95% confidence interval. P-values associated with each beta value were corrected for multiple testing using the FDR method (***p<0.001, **p<0.01, *p<0.05). The number of individuals with available data is shown.

**Extended Data Figure 10. Autism-OR among male and female individuals**. (**a**) For each autism-associated gene, the autism-OR among male individuals is compared to the autism-OR among female individuals. Some genes were not found mutated among either male or female autistic individuals. The gene-level autism-OR was measured using the sub-sampling procedure described in Methods, randomly selecting 1,596 individuals, i.e. the total number of female autistic individuals in the studied sample, for each sex and for each autism status 100 times. For genes on the X chromosome (highlighted in red), we selected genes with dominant mode of inheritance for female individuals (e.g. *MECP2*), and we did not filter for inheritance mode for male individuals.

## EXTENDED DATA TABLES LEGENDS

**Extended Data Table 1. List of the 185 autism-associated genes studied**. For each gene, the HGNC gene symbol, Entrez ID and ENSG ID are given, as well as their inclusion in the SPARK gene list, the SFARI score 1 list or the 102 TADA+ genes from Satterstrom et al. Cell 2020. We also provide the probability for loss-of-function intolerance score (pLI), the loss-of-function observed/expected upper bound fraction (LOEUF) and missense z-scores from gnomAD.

**Extended Data Table 2. List of the 2**,**492 constrained genes studied**. For each gene, the HGNC gene symbol, Entrez ID and ENSG ID are given, as well as the probability for loss-of-function intolerance score (pLI), the loss-of-function observed/expected upper bound fraction (LOEUF) and missense z-scores from gnomAD.

**Extended Data Table 3. Prevalence and autism-OR for S-LoFs in autism-associated genes**. For each autism-associated gene, the number of carriers among diagnosed and undiagnosed individuals, the autism-OR and empirical p-value with 95% CI are shown (Methods).

**Extended Data Table 4. Prevalence and autism-OR for S-LoFs in constrained genes**. For each constrained gene, the number of carriers among diagnosed and undiagnosed individuals, the autism-OR and empirical p-value with 95% CI are shown (Methods).

**Extended Data Table 5. Prevalence and autism-OR for S-SYNs in autism-associated genes**. For each autism-associated gene, the number of carriers among diagnosed and undiagnosed individuals, the autism-OR and empirical p-value with 95% CI are shown (Methods).

**Extended Data Table 6. Brain spatiotemporal gene expression levels**. Mean and standard deviation of the expression of all autism-associated genes across four brain regions and eight developmental periods are provided. Note that P1R4 is not listed, due to insufficient number of individuals in the BrainSpan study (Methods).

**Extended Data Table 7. Pearson correlation between autism-OR and brain spatiotemporal gene expression level for autism-associated genes**. For each brain region and each developmental period, the Pearson R and corresponding p-value is shown, along with corrected p-values using the FDR method. Note that P1R4 is not listed, due to insufficient number of individuals in the BrainSpan study (Methods).

**Extended Data Table 8. Pathway annotation of autism-associated genes**. Absence and presence of each autism-associated gene in lists of chosen annotations (SynGO and Gene Ontology Transcription, DNA-templated) is indicated.

**Extended Data Table 9. Autism-related and socio-economic traits regression results for S-LoFs in autism-associated genes, sex and autism-PGS**. For each response variable and each autism-OR threshold, the number of carriers and non-carriers among diagnosed and undiagnosed individuals, the standardized beta values and corresponding 95% confidence interval, the p-values and the corrected p-values are given for each covariate of the regression model. Corrected p-values based on the FDR method, independently for each covariate and separately for i) autism status, SCQ t-score, IQ score and autism factors, ii) developmental milestones and iii) socio-economic features and fluid intelligence. The number of carriers and non-carriers of S-LoFs in autism-associated genes are also shown.

**Extended Data Table 10. Autism-related and socio-economic traits regression results for S-LoFs in constrained genes, sex and autism-PGS**. For each response variable and each autism-OR threshold, the number of carriers and non-carriers among diagnosed and undiagnosed individuals, the standardized beta values and corresponding 95% confidence interval, the p-values and the corrected p-values are given for each covariate of the regression model. Corrected p-values based on the FDR method, independently for each covariate and separately for i) autism status, SCQ t-score, IQ score and autism factors, ii) developmental milestones and iii) socio-economic features and fluid intelligence. The number of carriers and non-carriers of S-LoFs in constrained genes are also shown.

**Extended Data Table 11. Autism status regression results for S-LoFs and autism-PGS in autism and constrained genes in the iPSYCH sample**. For each gene set and each autism-OR threshold, the standardized beta values and corresponding 95% confidence interval, p-values and corrected p-values are given for each covariate of the regression model. Corrected p-values based on the FDR method, separately for autism and constrained gene sets. The number of carriers and non-carriers of S-LoFs in autism and constrained genes are also shown.

**Extended Data Table 12. Brain anatomy regression results for the 1**,**940 carriers of S-LoFs in autism or constrained genes**. For each modality, region and hemisphere, the standardized beta values and corresponding 95% confidence interval, p-values and corrected p-values are given for autism-OR and sex. Corrected p-values based on the FDR method, independently for each covariate and separately for i) total values for global analyses (cortical thickness, cortical surface area, cortical volume), ii) each modality for region-specific analyses. The number of carriers of S-LoFs in autism or constrained genes are also shown.

**Extended Data Table 13. Odds ratio of participation for carriers of variants in autism-associated or constrained genes**. For each feature and variant/gene set, the number of carriers and non-carriers of variants among respondent and non-respondent individuals are shown, as well as the odds ratio for participation, its 95% confidence interval, p-value and corrected p-value.

