## Supplementary figures and images for "Sub-diagnostic effects of genetic variants associated with autism"

### Extended Data Figure 1

#### Extended Data Figure 1

##### SSC and SPARK admixture

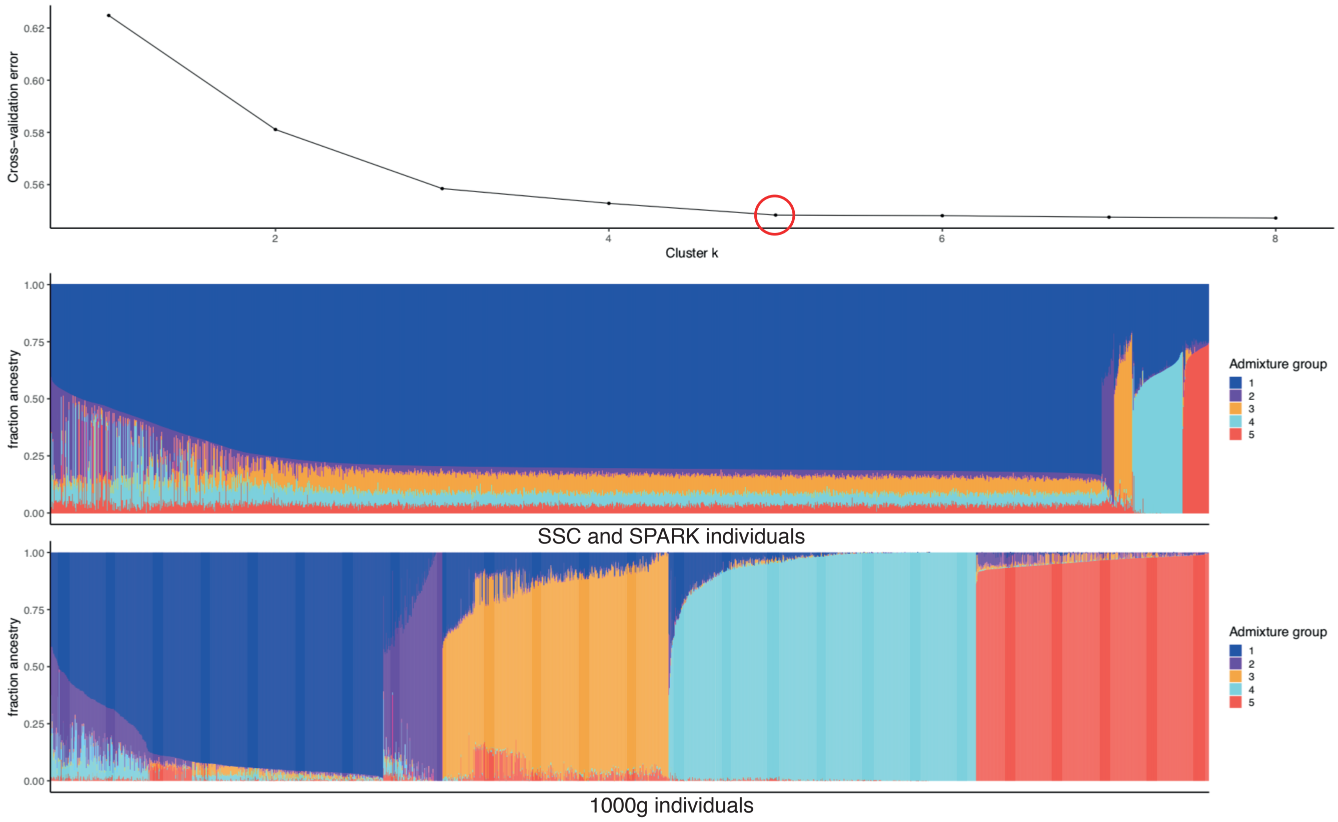


##### UK-Biobank admixture

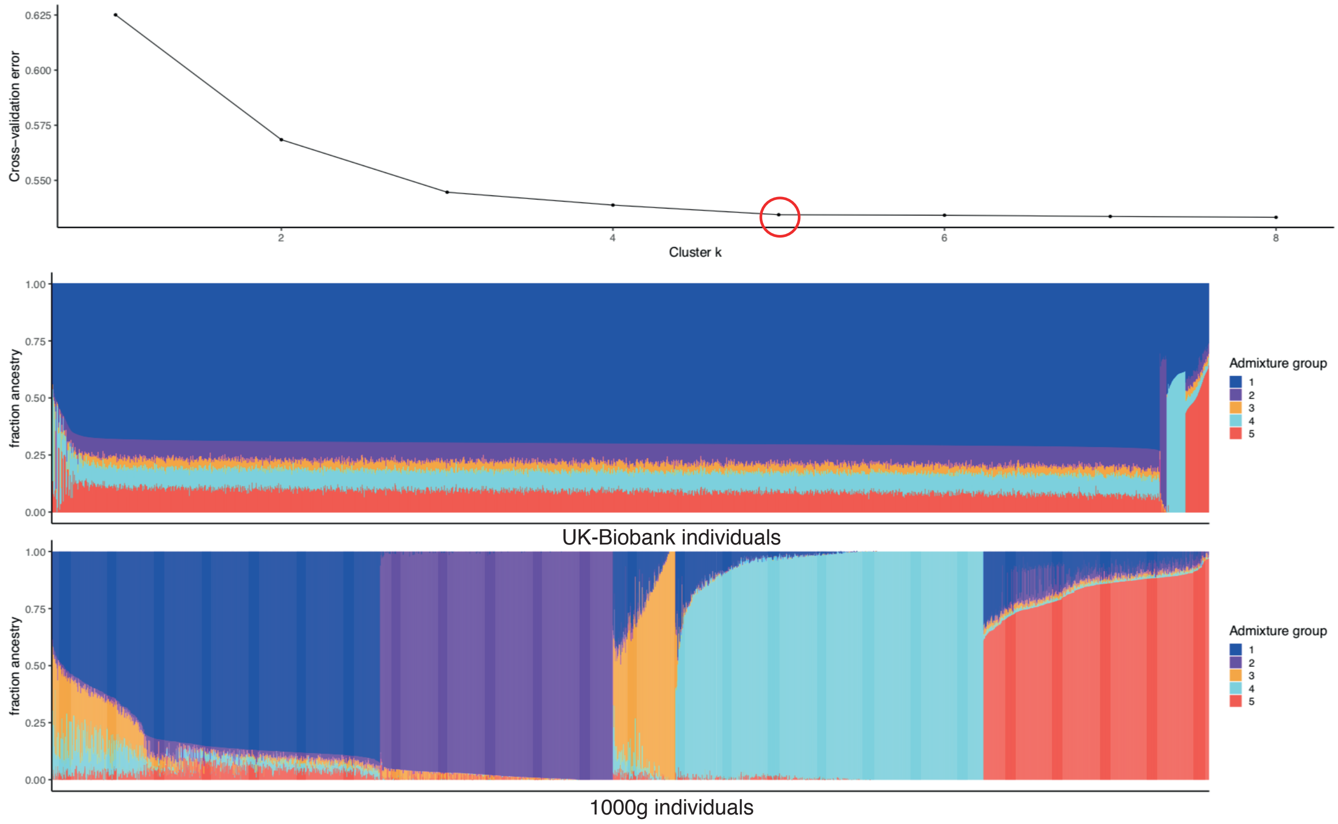

### Extended Data Figure 3

**Extended Data Figure 3**

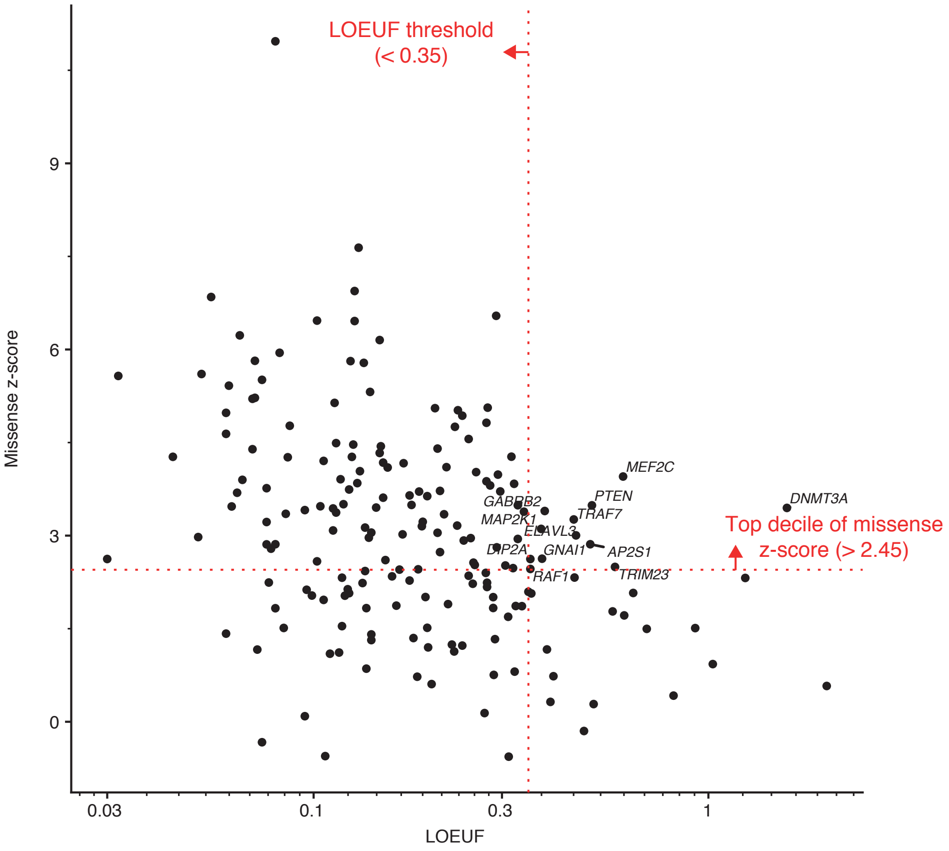


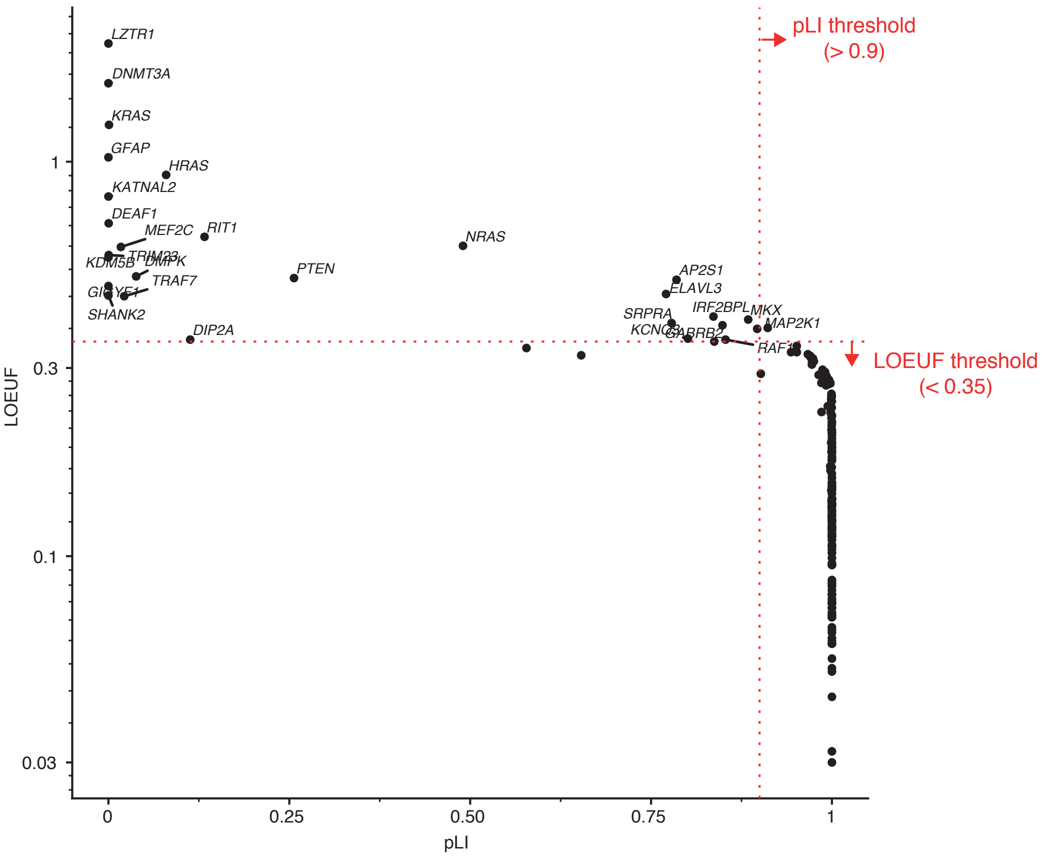

### Extended Data Figure 4

#### Extended Data Figure 4

##### *SHANK3*

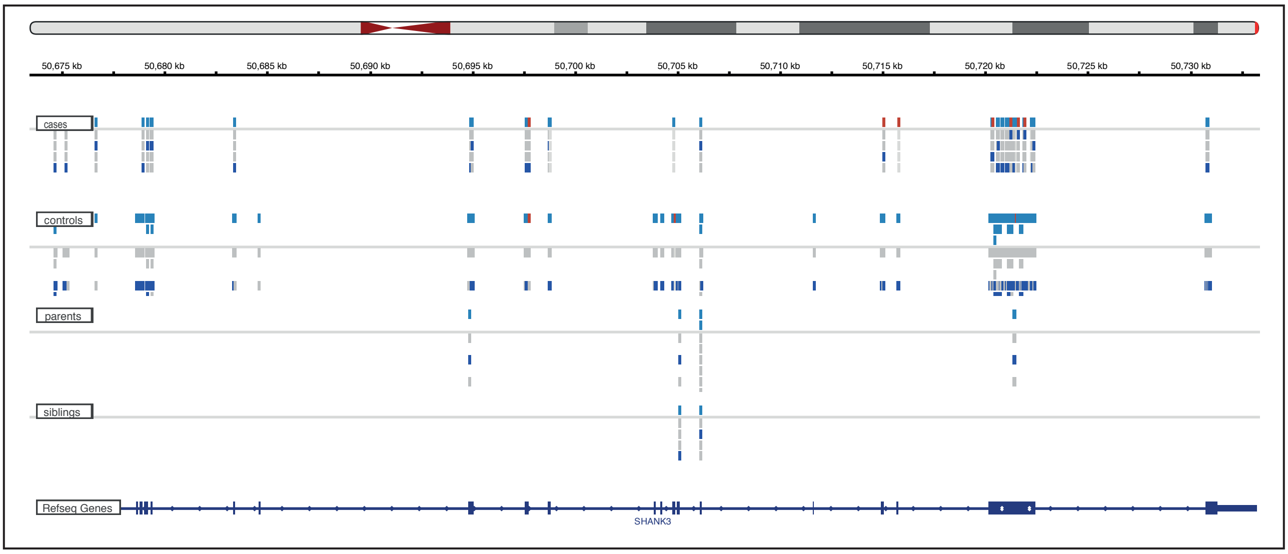


##### *NRXN2*

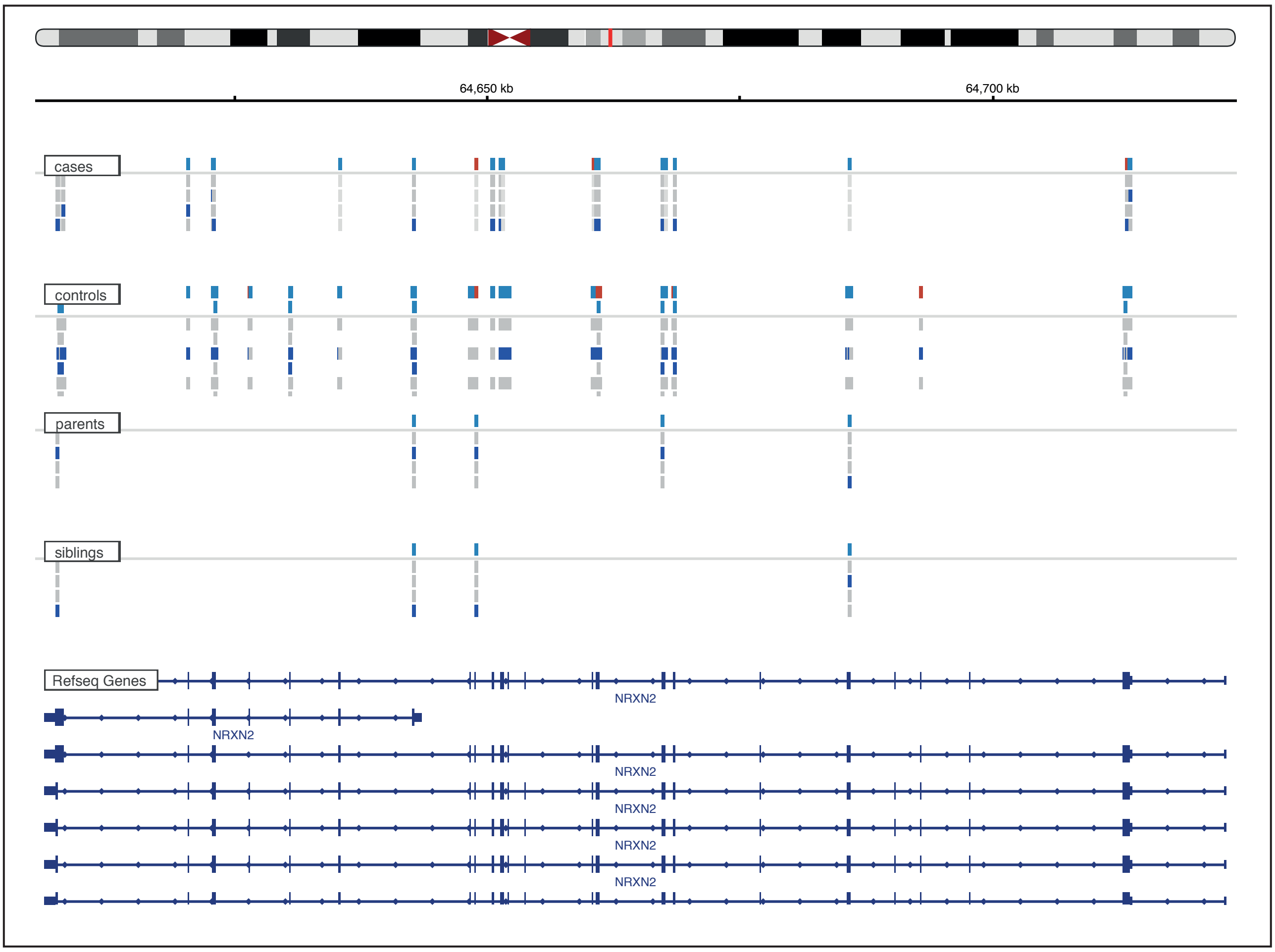

### Extended Data Figure 5

**Extended Data Figure 5**

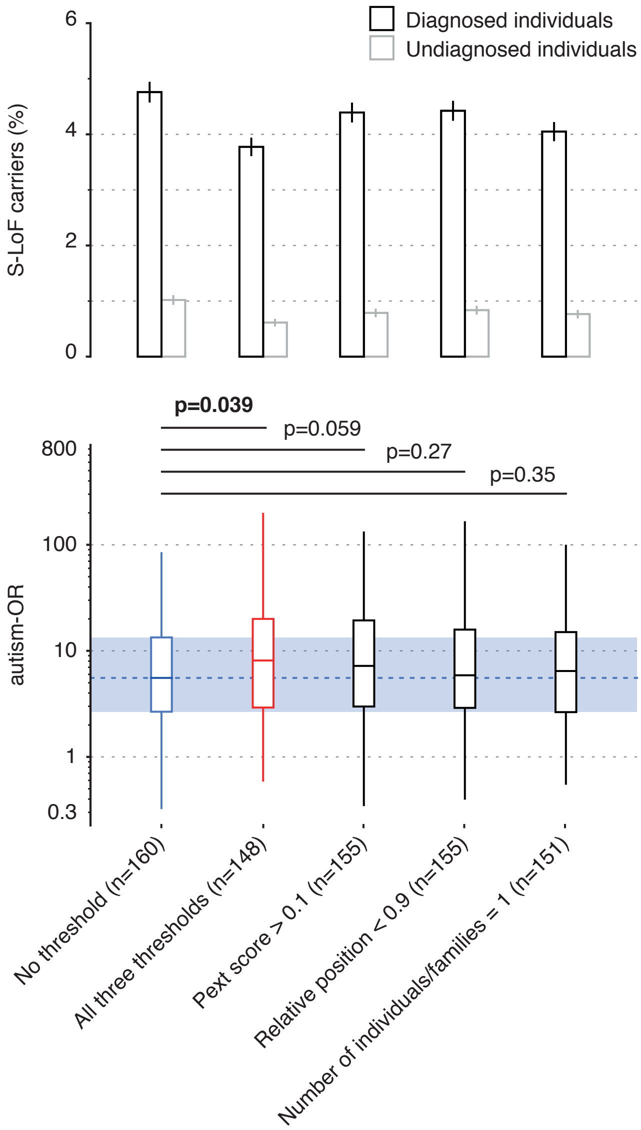

### Extended Data Figure 6

**Extended Data Figure 6**

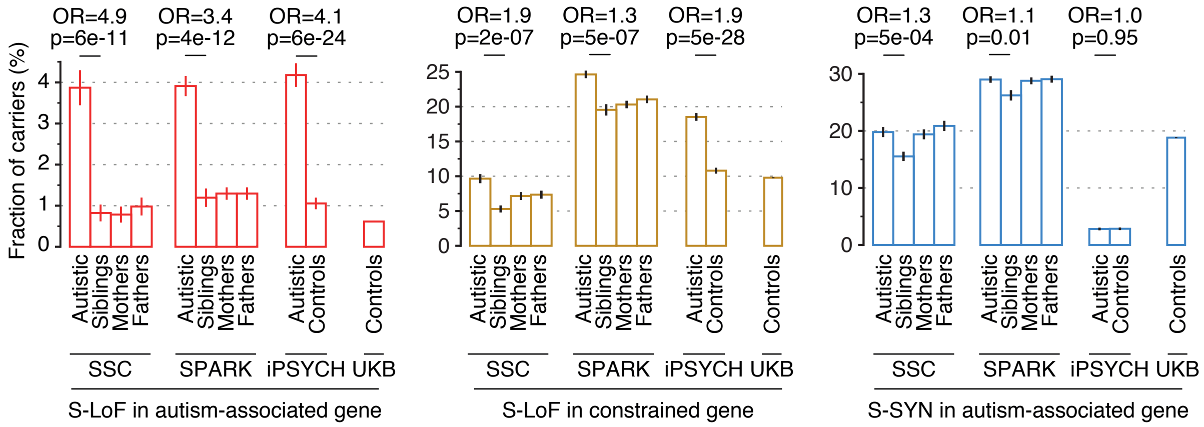

### Extended Data Figure 7

**Extended Data Figure 7**

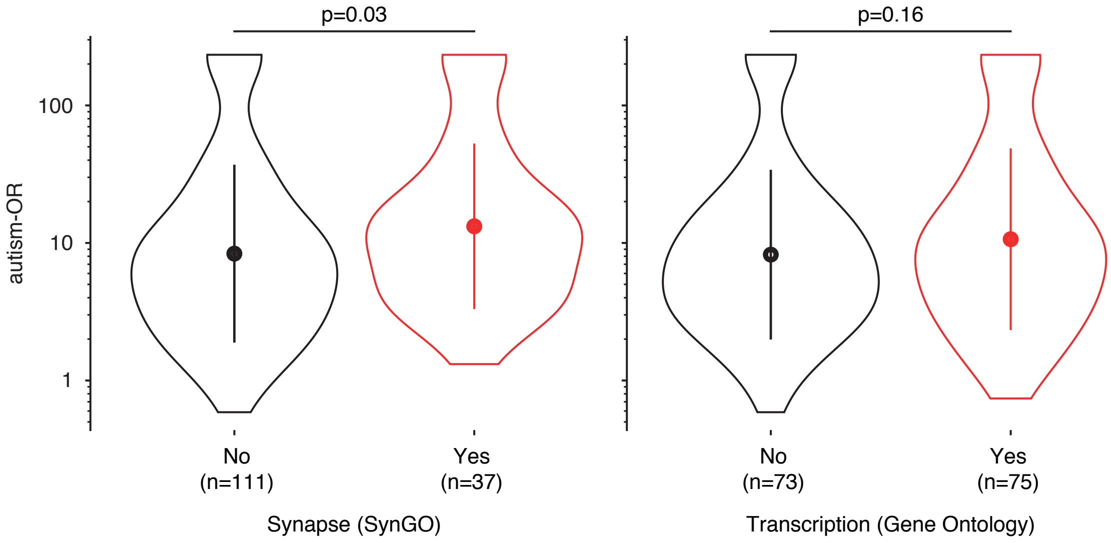

### Extended Data Figure 8

**Extended Data Figure 8**

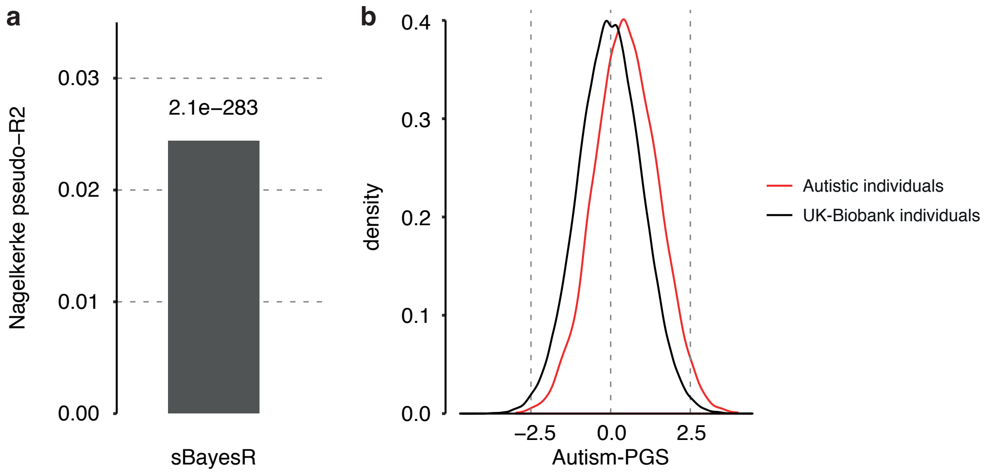

### Extended Data Figure 9

**Extended Data Figure 9**

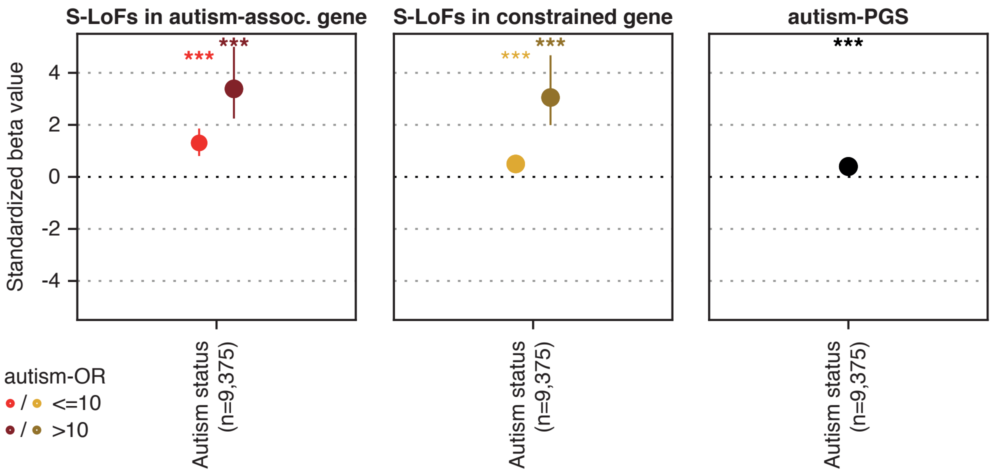

### Extended Data Figure 10

**Extended Data Figure 10**

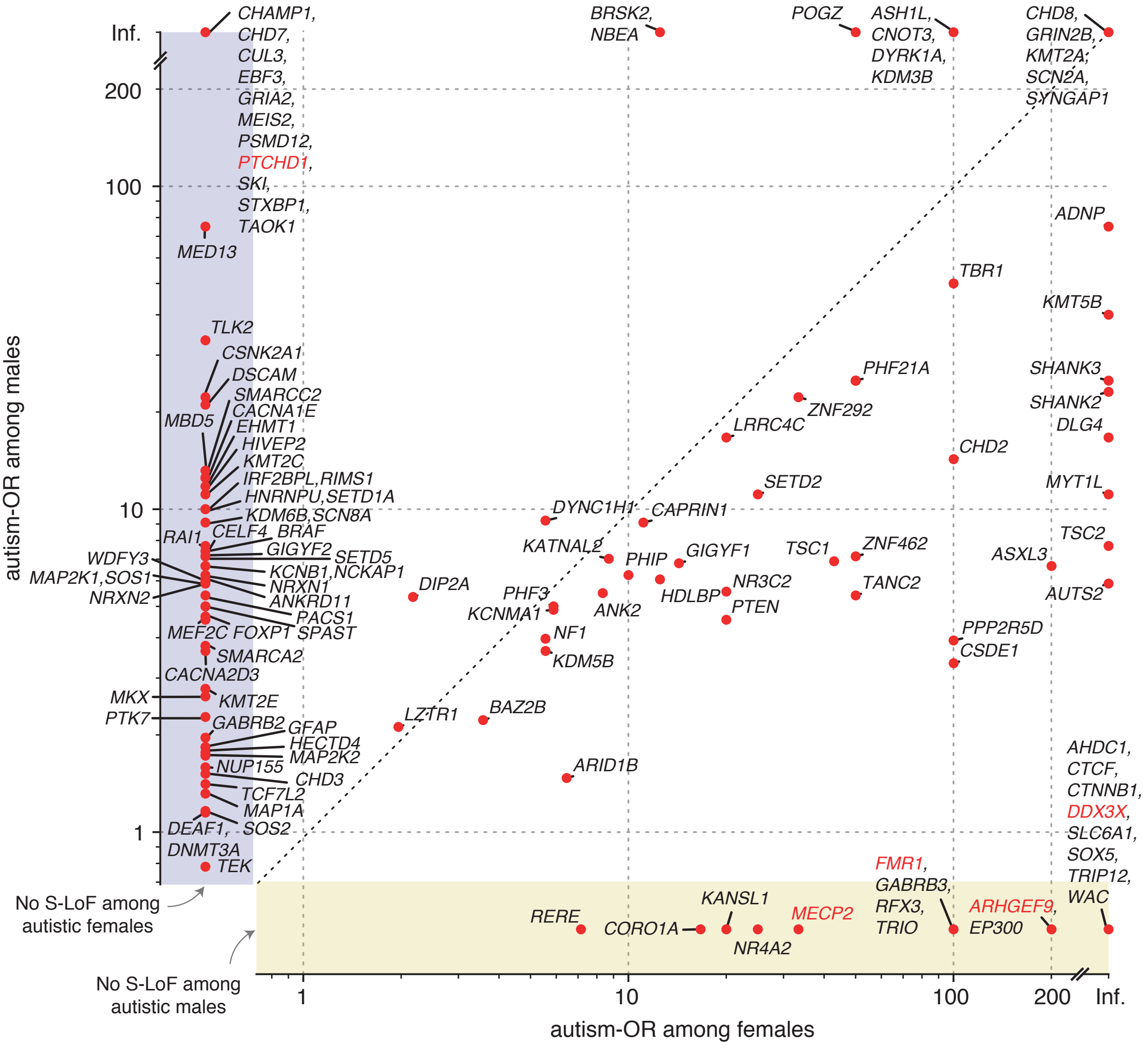
