## Extended Data Figure 2 for "Sub-diagnostic effects of genetic variants associated with autism"

**Meta-analysis $n_{ASD}$ = 13,091, $n_{relatives+controls}$ = 213,573**

| **Cohort** | **UK-Biobank** | **SSC** | **SPARK** | **iPSYCH** |
|---|---|---|---|---|
| Probands | - | 2,041 | 6,239 | 4,811 |
| Siblings | - | 1,944 | 2,344 | - |
| Parents | - | 4,082 | 11,118 | - |
| Controls | 188,871 | - | - | 5,214 |

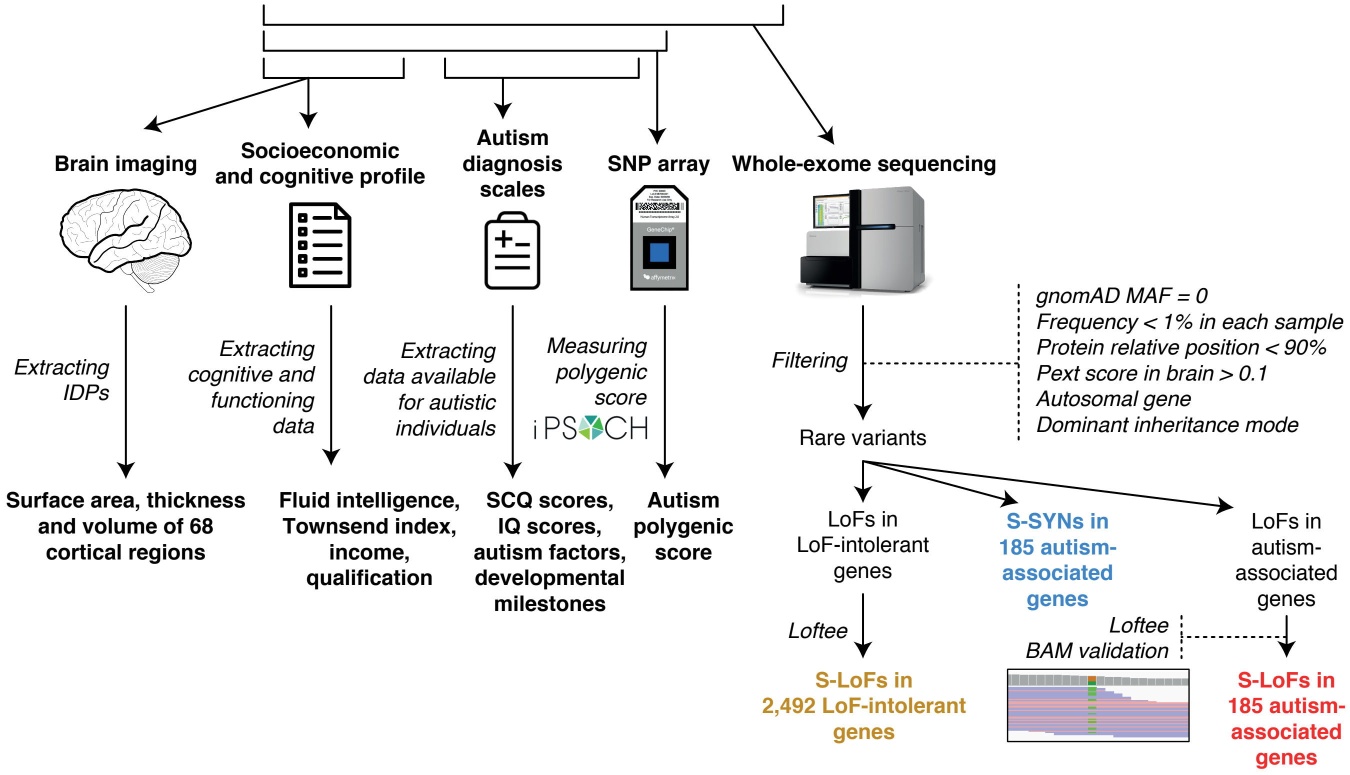
